# Bacterial and Fungal Co-Infections among ICU COVID-19 Hospitalized Patients in a Palestinian Hospital: Incidence and Antimicrobial Stewardship

**DOI:** 10.1101/2021.09.12.21263463

**Authors:** Hani A. Naseef, Ula Mohammad, Nimeh Al-Shami, Yousef Sahoury, Abdallah D. Abukhalil, Mohammad Farraj

## Abstract

Diagnosis of co-infections with multiple pathogens among hospitalized COVID-19 patients can be jointly challenging and very essential for appropriate treatment, shortening hospital stay and preventing antimicrobial resistance. This study proposes to investigate the burden of bacterial and fungal co-infections outcomes on COVID-19 patients. It is a single centre cross-sectional study of hospitalized COVID-19 patients at Beit-Jala hospital in Palestine. The study included 321 hospitalized patients admitted to the ICU between June 2020 and March 2021 aged ≥ 20 years, with a confirmed diagnosis of COVID-19 via RT-PCR conducted on a nasopharyngeal swab. The patient’s information was gathered using graded data forms from electronic medical reports. The diagnosis of bacterial and fungal infection was proved through the patient’s clinical presentation and positive blood or sputum culture results. All cases had received empirical antimicrobial therapy before the ICU admission, and different regimens during the ICU stay. The rate of bacterial co-infection was 51.1%, mainly from gram-negative isolates (Enterobacter species and K.pneumoniae). The rate of fungal co-infection caused by A.fumigatus was 48.9%, and the mortality rate was 8.1%. However, it is unclear if it had been attributed to SARS-CoV-2 or coincidental.

## Introduction

Severe Acute Respiratory Syndrome Coronavirus 2 (SAR-CoV-2) is a highly contagious novel viral pathogen that provokes an immediate spread among hospitalized patients with Community-Acquired Pneumonia (CAP) (1); extending from mild symptoms approximately in 84% of patients to life-threatening hypoxic conditions necessitating admittance into the Intensive Care Unit (ICU) and oxygen support (2). Additionally, it causes multi-organ failure that involves sepsis and thromboembolic complications, which progresses into an acute kidney and cardiac injury (3). On March 11^th^, 2020, the World Health Organization (WHO) deemed COVID-19 a pandemic disease; due to its unusual transmission speed and the wide-scale of infection (4).

The bacterial and fungal co-infections are frequently recurring due to respiratory viral diseases. For example, most deaths in the Spanish flu pandemic in 1918 were due to subsequent bacterial infection (5). It raises a concern as it hinders COVID-19 management, worsens prognosis, and might increase the fatality rate. The reported prevalence of bacterial co-infection varies between studies (6).

Throughout the pandemic, evaluation of gathered specimens from hospitalized COVID-19 patients reported the most prevalent organisms, which induce the co-bacterial infection; these were S. aureus, S. epidermidis, H. influenzae, Streptococcus spp., E. coli, K. pneumoniae and P. aeruginosa (7). Therefore, many COVID-19 treatment protocols include empirical antibiotic therapy to cover suspected organisms. Relatively, in a systematic review, thirty studies were summarized including 3834 COVID-19 patients. Overall only 7% of the hospitalized patients were confirmed to have bacterial co-infection with one or more pathogens, primarily in seriously ill patients (8). On the contrary, fungus infections in COVID-19 patients are neglected or delayed and were elusive to have the actual prevalence of fungal co-infection and yet to be explained in extent. Few studies were conducted in this regard; a systematic review meta-analysis research was carried out in China on 2780 confirmed SAR-CoV-2 participants from nine relevant articles. After fungal culturing at admission, 0.12%- 0.15% of the cases were positive for fungal infection and Asian patients were more likely to acquire fungal co-infection. The infecting fungi include Aspergillus, Candida, Cryptococcus neoformans, Pneumocystis, or other fungal species (9).

The exact mechanism of microbial co-infection is inadequately understood. Virologists assumed that the viruses attach and penetrate the host’s airway epithelial cells creating an inflammatory response desensitizing Toll-like receptors leading to cellular apoptosis in numerous mechanisms. In addition, it debilitates the body’s defense, induces cellular dysfunction and death (10).

Viruses aid in proliferation, colonization, and invasive infection of opportunistic nosocomial pathogen or respiratory tract normal flora; via hindering the innate immune response, compressing airway mucus, and disrupting the cilia. Thereby, this will allow for the spread and adhesion to more sites (3). Furthermore, various researches highlighted the synergy regulations among viruses and bacteria; both are jointly advantageous, aggravating clinical outcomes. If this cooperation is confirmed, then the management with antibiotics becomes further reasonable (11).

The COVID-19 disease has been chiefly linked to high levels of inflammatory markers, such as elevated levels of C-reactive protein and procalcitonin. Accordingly, the similarity between Sar-CoV-2 and bacterial infections in radiological infiltrates and laboratory findings makes it challenging in daily medical practice to provide a precise diagnosis (12). However, according to current studies, most COVID-19 patients did not have bacterial or fungal co-infections upon admission or through the hospital stay, even though they received antibiotics upon admission or before diagnosis (13). Furthermore, lack of specific antiviral agents and the overload to the health services capacity, saturated laboratories used to identify the causative microbial and diagnosis uncertainty to rule out super-infections; those factors advocated escalation of antimicrobial consumption in the pandemic (14). As a result, clinicians are obliged to prescribe broad-spectrum multi-antibiotic regimens; to treat all the possible infected pathogens. Therefore, this study aimed to investigate and evaluate bacterial and fungal co-infection burden and prevalence among COVID-19 patients admitted to a treatment facility at Beit-Jala hospital in Palestine. In addition, it reveals the etiology of the infection, provides the antimicrobial stewardship that was used in the hospital and gives insight into the association of clinical outcomes with several factors and comorbidities.

## Material and Methods

### Study Design and Study Population

This single-centre cross-sectional study was conducted on patients aged 20 years and older in Beit-Jala hospital, one of the central institutes for treating SAR-CoV-2 patients in Palestine. A total of 321 critically ill patients were admitted to the ICU between June 2020 and March 2021, with a confirmed diagnosis of COVID-19 via reverse transcriptase-polymerase chain reaction assay (RT-PCR) conducted on nasopharyngeal swab specimens. Only patients with positive cultures for bacteria or fungi were included in the study, and patients with negative cultures were excluded. The study was approved by the Scientific Research Ethics Committee of Birzeit University and by Beit-Jala hospital.

### Data Extraction and Analysis

Data extraction was done manually and verified by a second researcher, using graded data forms from electronic medical reports. The obtained data were socio-demographics, chronic comorbidities, laboratory findings (CRP, Leukocytes, blood Oxygen saturation), duration of ICU stay and other factors. The bacterial or fungal co-infection was proved through clinical presentation and positive blood or sputum testing via Laboratory Information System. Before and during the ICU admission, antimicrobial utilization was recorded, and the reports were double-checked for accuracy and completeness.

The analysis was performed on these data and summarized socio-demographic and clinical characteristics using relevant descriptive statistics; categorical variables as percentages and frequencies. Then Pearson’s Chi-square test was performed to determine the association between the main parameters, which were the ICU residency duration and the other factors such as; demographics, Iron supplements, antibiotics administration before and during the ICU admission and smoking habits, etc. Data was applied and presented upon the IBM Statistical Package for the Social Sciences version 22.0. (SPSS).

### Definitions

Co-infection is a sequela that occurs during or after the primary causative pathogen or its treatment. The isolated bacteria or fungi obtained from COVID-19 patients should be clinically relevant and not microbiota or contaminants. All the microbiological samplings were conducted upon the hospital’s entry and before the ICU admittance, and only patients holding positive bacterial or fungal outcomes were admitted into the unit. In combination with decision-making, a scope of standards should meet to provide permission on who necessitates access. Essentially, Computerized Tomography (CT) scans revealing ground-glass opacification, which correlated with Pneumonia. In addition, supplementary laboratory outcomes were crucial in making the diagnosis, such as C-reactive protein (CRP) level, Oxygen saturation (SpO2), Leukocytes numbers and body temperature. Patients were immediately admitted to the ICU; if two or more of the above matched positive isolates test and CT-scan showing Pneumonia.

To determine the status of each patient, laboratory findings must be evaluated. For example, CRP values that ranged between 4.5-8.5 mg/dL were rated as moderate and ≥ 8.5 mg/dL and then rated as severe. Leukocytes results ranged within 4.6-8 g/L was considered mild, while >8 g/L indicates seriously ill patients. Normal Oxygen saturation of ≥ 95%, but in COVID-19 patients, the oximeter reading at rest of 90% or lower is a severe case. The body temperature is deemed high if it was 38-39 °C and very high if it was >39 °C.

## Results

### Presenting Demographics

A total of 321 moderate-severe ill patients were screened and evaluated. The patient’s demographics and clinical characteristics are presented in Table 1. Overall, 48.9% (170) were 41-64 years of age, 50.5% (162) were males. Prevailing chronic medical conditions included Diabetes Mellitus 77.9% (250), Atrial Hypertension 66.4% (213), Obesity 36.8% (118), Cancer 20.6% (66), Coronary Heart Disease 23.1% (74) and 3.7% of patients did not have personal clinical history. 58.6% were not smokers, and 62.6 % were supplemented with Iron 376 mg/day intravenously for three days. Further, all the participants in this study had received N-acetyl-cystine 1200 mg /day, which began from their hospital’s entrance to the exit.

**Table 1:**
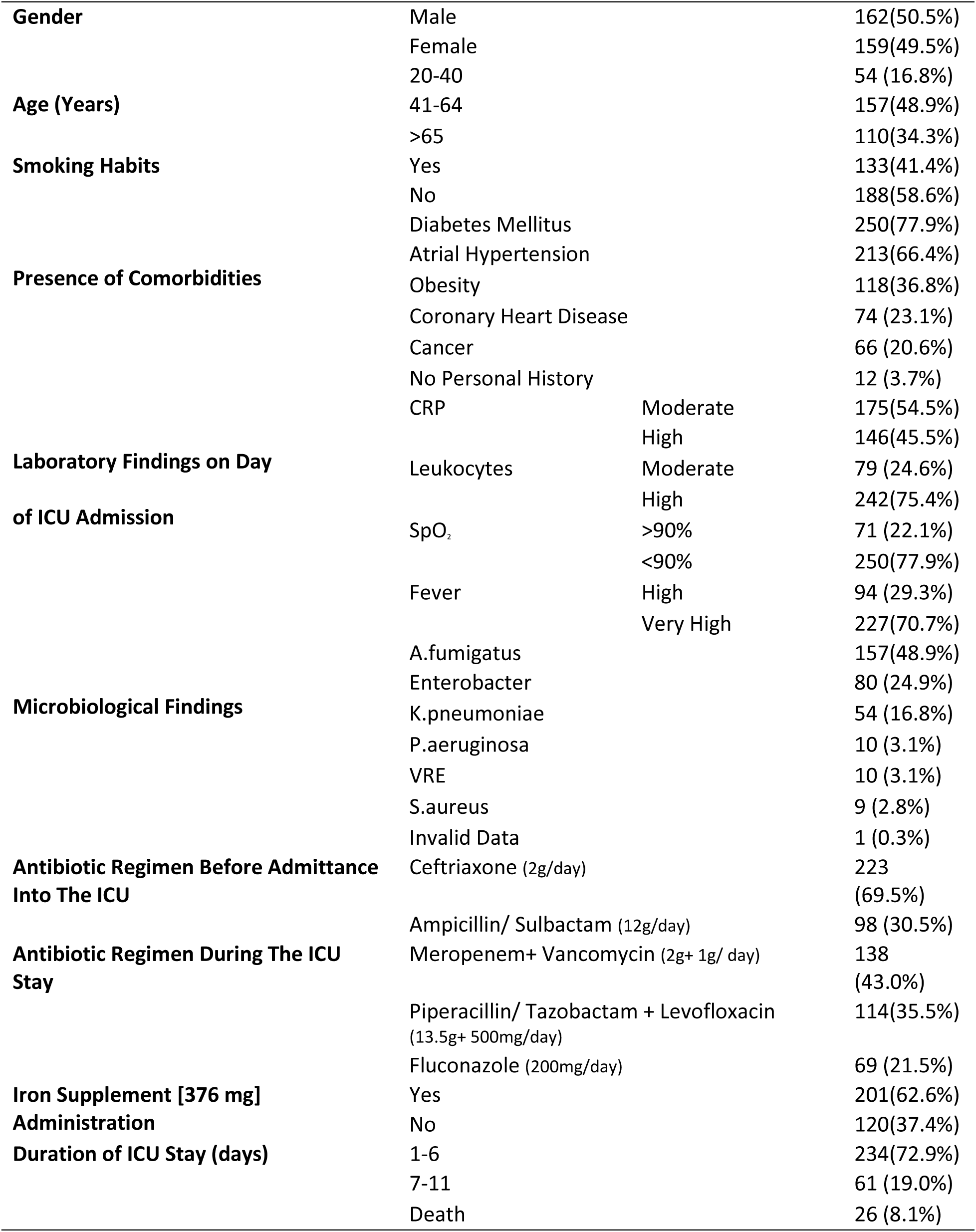
Demographics, Medical History and Antibiotic Regimens. (N 321)

### Bacterial and Fungal Co-Infection

The overall mortality rate in this study was 8.1% (26), while 72.9% (234) of the patients remained in the ICU for 1-6 days. Each patient was infected with one type of bacteria or fungi; presented as respiratory infection, bacteremia, and skin infections. A total of six different pathogens were detected in the samplings and considered as the infecting organisms. The most abundant types were Aspergillus fumigatus 48.9% and Enterobacter 24.9%, as shown in Table 1

### Patterns of Antibiotic Use

In terms of antibiotic prescription patterns, through the first 24h of hospital admission and based on per case specificity, the proper liver or kidney function tests had been conducted in all cases; to determine the appropriate regimen for bacterial or fungal infections. Before the ICU access, the treatment span was six days on average; 69.5% (223) received Ceftriaxone, and 30.5% (98) patients had received Ampicillin/ Sulbactam; both regimens were administered as intravenous dosing. Upon ICU admission, every patient had been given one kind of IV antibiotic regimen throughout the stay; 43.0% (138) took Meropenem and Vancomycin and 35.5% (114) given Piperacillin/ Tazobactam and Levofloxacin. By January and after getting financial support from the Ministry of Health, patients infected with A.fumigatus started therapy with Fluconazole, 21.5% (69) were successfully treated. Considering different regimens had notable distinct outcomes, analysis was performed to determine the association linking antimicrobial therapy class and the ICU stay or death (Table2). After hospital discharge, patients taking antibacterial medications are switched to oral Azithromycin. Patients treated with Fluconazole IV for two days switched to oral Fluconazole 150mg/ week.

**Table 2:** Impact of Different Non-Significant Values on The duration in the ICU. (N 321)

| Parameter |  | Duration 1-6<br>Days | Duration 7-11<br>Days | Death | P value |
| --- | --- | --- | --- | --- | --- |
| **Age**<br>(years) | **20-40** | 41(75.9%)* | 10(18.5%) | 3(5.6%) | 0.413 |
|  | **41-64** | 119(75.8%) | 28(17.8%) | 10(6.4%) |  |
|  | **>65** | 74(67.3%) | 23(20.9%) | 13(11.8%) |  |
| **Diabetes** | **Yes** | 180(72.0%) | 49(19.6%) | 21(8.4%) | 0.794 |
|  | **No** | 54(76.1%) | 12(16.9%) | 5(7.0%) |  |
| **Hypertension** | **Yes** | 156(73.2%) | 43(20.2%) | 14(6.6%) | 0.317 |
|  | **No** | 78(72.2%) | 18(16.7%) | 12(11.1%) |  |
| **Obesity** | **Yes** | 81(68.6%) | 25(21.2%) | 12(10.2%) | 0.383 |
|  | **No** | 153(75.4%) | 36(17.7%) | 14(6.9%) |  |
| **Smoking** | **Yes** | 103(77.4%) | 25(18.8%) | 5(3.8%) | 0.052 |
|  | **No** | 131(69.7%) | 36(19.1%) | 21(11.2%) |  |
| **CRP** | **Moderate** | 128 (73.1%) | 33 (18.9%) | 14 (8.0%) | 0.994 |
|  | **High** | 106 (72.6%) | 28 (19.2%) | 12 (8.2%) |  |
| **Leukocytosis** | **Moderate** | 59 (74.7%) | 18 (22.8%) | 2 (2.5%) | 0.089 |
|  | **High** | 175 (72.3%) | 43 (17.8%) | 24 (9.9%) |  |
| **SpO2** | **<90** | 188 (75.2%) | 40 (16.0%) | 22 (8.8%) | 0.032 |
|  | **>90** | 46 (64.8%) | 21 (29.6%) | 4 (5.6%) |  |
| **Fever** | **High** | 71 (75.5%) | 19 (20.2%) | 4 (4.3%) | 0.265 |
|  | **Very High** | 163 (71.8%) | 42 (18.5%) | 22 (9.7%) |  |

\*The percentages indicate proportion of patients within each category in any duration.

Table 2 reported that there was no significant association between the ICU stay duration with age, laboratory test results or having a chronic disease. A significant association found between the residency duration and SpO2 (P-value= 0.032), that patients whose SpO2 levels were <90% (188:75.2%) were stayed for a shorter duration (1-6 days) at the ICU compared to patients with SpO2 reads >90% (46: 64.8%).

Table 3 showed a significant association between the ICU residency duration and the infecting isolates (P-value= 0.008). The patients suffering from fungal co-infection 126 (80.3%) stayed fewer days in the ICU comparing with patients with bacterial co-infection 108 (65.9%).

**Table 3:** Impact of Different Significant Values on The duration in the ICU.

| Parameter | Duration 1-6 Days | Duration 7-11 Days | Death | P |
| --- | --- | --- | --- | --- |
| **Ampicillin/Sulbactam** | 52 (53.1%) ** | 29 (29.6%) | 17(17.3%) |  |
| **Ceftriaxone** | 182 (81.6%) | 32 (14.3%) | 9 (4.0%) |  |
|  |  |  |  | <.001 |
| **Piperacillin/Tazobactam+**<br>**Levofloxacin** | 57 (50.0%) | 37 (32.5%) | 20(17.5%) |  |
| **Meropenem+ Vancomycin** | 108 (78.3%) | 24 (39.3%) | 6 (4.3%) |  |
| **Fluconazole** | 69 (100%) | 0 (0%) | 0 (0%) | <.001 |
| **Yes** | 199 (99%) | 0 (0%) | 2 (1.0%) |  |
| **Iron Supplement No** | 35 (29.2%) | 61 (50.8%) | 24(20.0%) | <.001 |
| **Fungi** | 126 (80.3%) | 24 (15.3%) | 7 (4.5%) |  |
| **Bacteria** | 108 (65.9%) | 37 (22.6%) | 19(11.6%) | .008 |

**\*\* The percentages indicate proportion of patients within each category in any duration.**

Regarding therapy plan, there was a significant association between ICU residency duration and the empirical antibiotics (P-value <0.001), patients administered ceftriaxone 182(81.6%) were more likely to stay less at ICU compared with patients administered Ampicillin/Sulbactam 52 (53.1%). Patients who took fluconazole 69 (100%) were significantly more likely to stay less in the ICU compared to those administered Piperacillin and Tazobactam with Levofloxacin or Meropenem with Vancomycin 57 (50%), 108 (78.3%) respectively with P-value <0.001.

Moreover, patients administered iron supplements as part of their therapy regimen 199 (99%) were significantly more likely to stay for a shorter duration of time compared with those who did not administer iron 35 (29.2%, P-value <0.001).

## Discussion

In this research, the characteristics of co-infection in 321 severely ill COVID-19 patients were evaluated with a fatality rate of 8.1% in the ICU, compared to 25.7% mortality rate; that was obtained from a systematic review of the emerging literature including 15 studies carried out in countries most affected by the pandemic (16). Despite the absence of antimicrobial stewardship in urgent response, the hospital’s initiation of drastic empirical administration was crucial for either prevention or treatment of suspected infections. Hospital’s guideline adherence for empiric antimicrobial therapy and the other drugs was effective.

COVID-19 patients are at risk of fungal infections during the latter stages of the disease; due to seriously damaged alveoli and the decline in leukocyte counts (17). Based on the knowledge of SARS in 2003 and the invasive Aspergillosis, it is vital to consider the possibility of accompanied COVID and life-threatening fungal infection. At the beginning of blood/sputum culturing, the number of cases infected by A.fumigatus was 157. Initially, those patients were given empirical antibacterial regimens and in the ICU; because it was costly to be obtained by the hospital. By January, confirmed fungal infections were treated with Fluconazole. This had reduced the average ICU stay from six days to less than two days, and the mortality rate became 0% rather than 4.5% due to A.fumigatus invasive disease. Throughout the pandemic, specialists realized the risks of fungal co-infection. Therefore, the French High Council for Public Health recommended that clinicians should concentrate on fungal infections in COVID patients, particularly in severe cases of the disease.

Bacterial co-infection was recorded in 51.1% of cases, with higher mortality rates among Enterobacter infected patients; this may be due to its known drug resistance. It should be noted that normal oral flora colonization or contamination may occur, particularly in the sputum specimens. In general, the empirical treatment with Ceftriaxone or Ampicillin/Sulbactam showed minimal benefits to the clinical status. However, we observed a better prognosis with Ceftriaxone than Ampicillin/Sulbactam. The mortality rate also varied from 4% with Ceftriaxone vs. 17.3% with Ampicillin/Sulbactam; this association needs more investigation.

Once the bacterial co-infection is confirmed and the patient is admitted into the ICU, the wide-spectrum antimicrobial regimens are administered. Unfortunately, there is no available data to compare COVID-19 patients who received antibacterial agents with those who did not; to determine the therapeutic efficacy. However, Piperacillin with Tazobactam and Levofloxacin resemble Meropenem and Vancomycin in the clinical outcomes and management rate. Counter to a study that reveals a higher rate of mortality and organ failure regarding Meropenem (13). Furthermore, after the recovery and discharge from the hospital, all outpatients have been prescribed Azithromycin, which also has antiviral activity and may decrease the incidence of acute organ failure (13).

Earlier examinations illustrated that multiple antibiotic administrations did not alter the disease results or had joined with higher deaths (18). In our study, it appeared to induce a good prognosis. Ceftriaxone, Meropenem, and vancomycin regimens have been linked with more favorable outcomes and lower mortality rates. Until solid evidence is available, antimicrobials should be maintained for critical cases and constantly re-evaluated based on the patient’s progress. Most importantly, the therapy must only cover the suspected or confirmed bacterial infection.

Antibiotic resistance and allergic reactions are multifactorial issues, and the pandemic has frightened the health system and stimulated pressure on bacteria. In our study, all COVID-19 patients were admitted to the ICU and received at least four diverse types of broad-spectrum antibacterial agents; in addition, the accessibility to over-the-counter antimicrobials had undeniably grown through the pandemic. This inappropriate behavior will lead to the evolution of high levels of resistant bacteria.

The unusual incidence of bacterial or fungal infections in our study differs from other published studies (9) (19). This significant finding is due to the participant characteristics of critically ill patients receiving therapy via invasive catheters and being at risk of infection by nosocomial pathogens. Moreover, the unusual rate of infection had been credited to the exhaustion in the clinical care system creating a prolonged hospital exposure, principally in the major waves of the virus.

In our study, 100% of the patients were severe-critical ill. Therefore, it is required to differentiate between them and mild-moderate patients and prioritize the ICU admittance. Fever was the notable symptom in SARS-CoV-2 infection identified at the early stages of the disease. Regarding laboratory findings, leukocytosis occurred in critically ill patients with distinct grades, and it was observed in 11.4% severe COVID-19 cases (20); it aids in the disease progress evaluation. The intensity of leukocytosis reflects the strength of COVID-19 infection; it is exacerbated by the cytokine storm and inflammatory mediators that drive apoptosis and pulmonary microvascular destruction, creating alveolar oedema. The increased level of CRP is a diagnostic biomarker for coronavirus in distinguishing moderate and critical patients. The pathogenesis of COVID-19 involves a vigorous inflammatory response that manages a dysregulated flood of immune cells and signaling molecules (21). In general, viral respiratory infections have been linked to hypoxia, but COVID-19 individually starts significantly low Oxygen saturation levels (<94%), while the normal SpO2 is ≥ 95% at rest in healthy people. Hypoxia is a fearful condition that is monitored and adjusted by mechanical ventilation (22). Although several studies reported relations linking Laboratory findings and clinical outcomes, our cases did not show a significant link.

Throughout the coronavirus pandemic, the issue of tobacco smoking and the risk of infection were constantly reviewed. Published research indicated that smokers are twice as likely to develop severe COVID-19 as nonsmokers. Tobacco smoking induces alterations in the respiratory tract and cell-mediated immune response (23). Furthermore, current smokers have higher ace-2 gene expression than non-smokers; this gene is responsible for ACE-2 receptors production, which COVID-19 attaches to and penetrates the cells, thus raising the chance of infection (24). Our study did not find an association between smoking habits and the duration of ICU stay or clinical prognosis.

Numerous investigations found that older COVID-19 patients aged ≥ 50 years were correlated with higher risk for severe signs, atypical presentation, the opportunity for co-infections and higher fatality rates compared to patients with <50 years of age, and the rate usually increases rapidly with age; it is not surprising due to the drop in natural immunity by age, associated comorbidities and it is believed that older people are prone to adverse drug reactions. In general, people of different ages are prone to get infected with the COVID virus (25) (26). A review reported a twice higher fatality rate in males compared to females; it has stated that male patients may have higher ACE-2 enzyme activity, directed by male sex hormones, contributing to more risk for SARS-CoV-2 infection and worsening clinical outcomes (27). Additionally, the smoking rate is higher in males.

Effects of comorbidities such as hypertension, diabetes mellitus, coronary heart disease and Cancer were compared between survivors and non-survivors. All of those comorbid conditions have a significantly greater fatality rate in non-survivors; because they have decreased natural immunity and are poly-pharmacy patients (28) (29). In our study, there was no association between age, gender or comorbidities with the clinical outcomes and ICU stay duration.

Previous reports had described mild anemia in COVID-19 patients at the ICU; due to severe inflammation which consumes iron (30), the innate immune system limits the bioavailability of iron; to diminish viral replication and infection. Accordingly, iron absorption from the diet is reduced, and the liver generates hepcidin which blocks carrying iron out of the cell. Accordingly, iron supplementation may exacerbate the disease and increase the inflammatory process (31). However, no solid investigations were performed on the relation between iron and clinical results in COVID-19 patients. In our study, 62.6% of the patients had received IV iron supplement for three days, based on a randomized clinical trials, and 99% of them had improved and stayed for a shorter time (1-6 days) in the ICU; which provided a good immunity enhancing. Therefore, there is a strong association between low serum levels of iron and an increased infection rate and morbidity.

COVID-19 produces oxidative stress irregularity, and this process has been enhanced by glutathione reduction. Thiones are synthesized in a multi-step process in which N-acetyl-cysteine is part of it. Thiones are also ACE-2 blockers, thereby diminishing SARS-CoV-2 penetration into the cell. In a placebo-controlled study, NAC administration had reduced plasma inflammatory biomarker levels via several mechanisms. Besides antioxidant activity, NAC has vasodilator activity, especially in the intravenous route. In addition, it has mucolytic properties, inhibits RNA virus replication, and has confirmed protective effects against comorbid conditions, including cardiovascular diseases (32). In our study, patients had received NAC 1200 mg/day during the hospital stay, and even after discharge, those showed a better prognosis and improvement in their health status.

This research has some limitations: first, it is a cross-sectional study holding obstacles restricting the potential bias and difficulty proofing the relation between the exposure and the outcome. Other, it is a single-centre, and its results could vary in another setting. Comprehensively, those conclusions present data supporting a conservative antibiotic administration for severely unwell COVID-19 infected patients. Our examination regarding the impacts of employing antifungals to manage COVID-19 patients can work as a successful reference for future COVID-19 therapy.

## Conclusion

The COVID-19 pandemic exerts pressure on healthcare professionals, including a high probability of infection. It is eminent to save lives during this pressure, even if it means multi-drug antimicrobial regimen usages for prevention or treatment. However, the insufficiency of data about antibiotic types and dosage, and duration of treatment through the pandemic continue to be challenging to health care providers. Bacterial and fungal co-infection is common among COVID-19 patients at the ICU in Palestine; it is not evident if these cases are attributed to SARS-CoV-2 or coincidental; little of available data. Finally, Antimicrobial stewardship, appropriate patient assessment, and selecting the appropriate antimicrobial agents for the right patient at the right time will decrease hospital stay, mortality, and health care costs.

## Data Availability

The data used to support the findings of this study are available from the corresponding author upon request

## Acknowledgements

We would like to express our appreciation to the health worker at Beit-Jala Governmental Hospital, Palestine, for their support and cooperation.

## Ethics approval and consent to participate

Ethical approval of the study was obtained from Birzeit University. The permission to use the generated data was obtained from Beit Jala Governmental Hospital laboratory administration.

## Consent for publication

All authors read and approved the final manuscript

